# Identification of *HLA-A*, *HLA-B* and *HLA-C* triple homozygous and double homozygous donors: a path towards synthetic superdonor Advanced Therapeutic Medicinal Products

**DOI:** 10.1101/2025.05.19.25327154

**Authors:** Daniel Naumovas, Barbara Rojas-Araya, Catalina M. Polanco, Victor Andrade, Rita Čekauskienė, Beatričė Valatkaitė-Rakštienė, Inga Laurinaitytė, Artūras Jakubauskas, Mindaugas Stoškus, Laimonas Griškevičius, Ivan Nalvarte, Jose Inzunza, Daiva Baltriukienė, Jonathan Arias

## Abstract

Immune matching and rejection pose major hurdles in tissue transplantation. Here, we profile *HLA-A*, *HLA-B*, and *HLA-C* alleles in 3,496 Lithuanian donors genotyped at three-field resolution. The five most frequent alleles constitute 74.6% of *HLA-A*, 43.2% of *HLA-B*, and 59.2% of *HLA-C*, with HLA-A*02:01:01, HLA-B*07:02:01, and HLA-C*07:02:01 being the most common. Lithuanian allele frequencies closely resemble those of populations with pre-Neolithic hunter-gatherer ancestry, such as European-American and British groups. We identified 153 double homozygotes and 51 triple homozygotes for *HLA-A*, *HLA-B*, and *HLA-C*. Compatibility modeling showed triple homozygous profiles match 60.5% of Lithuanians (33.3% for double homozygotes), 13.4% of British population, and 7.4% of European-Americans. CRISPR-Cas9 guide RNA design yielded 54 candidates predicted to disrupt *HLA-A* or *HLA-B*, while preserving *HLA-C*, producing edited profiles matching over 98.1% of Lithuanians, 95.8% of European-Americans, and 95.6% of British population. Finally, we established 16 fibroblast lines from double and triple homozygotes, offering a resource for immune-compatibility studies and regenerative medicine applications.

## 2. Introduction

Transplantation of allogeneic organs, tissues and cells are constrained by immune-matching between the graft and the host. Immune matching is mediated by the human leukocyte antigen (HLA) genes. They are clustered in a 3.7M bp locus on chromosome 6, highly polymorphic and their inheritance reported as having intermediate linkage disequilibrium ^1,2^. Recent adoption in clinic of high resolution haplotyping, have improved the accuracy of immune matching for the more than 38,000 HLA alleles cataloged in the IPD-IMGT/HLA Database ^3^. Pursuing high level of matching is intended to minimize adverse events, such as graft-versus-host disease (GVHD) or immune rejection, which are frequently managed with immune suppressive drugs. There is a broad assortment of immune suppressive treatments for the management of transplantation, encompassing small molecule inhibitors, antimetabolites, corticosteroids, and antibodies ^4–6^. However, immune suppressive therapies are linked to an increased risk of infection ^7,8^. Therefore, pursuing high level of matching is intended to minimize adverse events caused by immune rejection and immune suppression. The importance of a high degree of immune matching for improving survival rates is well documented in the literature ^9^. Indeed, high levels of matching, referred as a complete HLA match ^10^, has a positive impact on patient survival.

HLA class I homozygous individuals offer increased immune compatibility with a relatively larger base of the population. They are individuals very scarcely represented in the population as expected from mendelian ratios. Cells from naturally occurring triple and double homozygous individuals are very valuable for the study of immune compatibility and applications of regenerative medicine. Genome editing tools are currently used to engineer synthetic immune compatibility, also called hypo-immunogenicity, in tissues and cells. This aid overcoming the challenges of identifying rare haplotypes in donor pools. Several approaches have been developed to bypass immune recognition by cytotoxic T cells while retaining self-recognition mediated by NK cells. The most frequent strategies through loss-of-function include the knock-out of specific HLA class I ^11^ and class II genes, beta-2-microglobulin (*B2M*) ^12,13^, *CIITA* ^14^, *TAP1* or *TAP2* and *CD74* ^15^. Conversely, the most frequent gain-of-function strategies involve the knock-in of *CD47* and *HLA-E* ^16^. Pioneering studies have demonstrated that gene editing depletion of *HLA-A* and *HLA-B* genes preserves the host NK cell recognition while preventing the CD8 T-cell mediated host-versus-graft rejection ^17^. This approach yields cells currently known as *HLA-C* retained. Triple and double homozygous samples are the ideal cell source for modulating immunogenicity, as they start from a relatively higher level of immune compatibility. Furthermore, they can be engineered in their *HLA* genes using programmable nucleases through simpler strategies compared to heterozygous samples. In this study we identify a cohort of naturally occurring triple and double homozygous individuals and isolated primary samples for prospective regenerative medicine applications. Additionally, we conducted an analysis of the frequency of class I HLA genes in the European-Lithuanian population, specifically characterizing the *HLA-A*, *HLA-B* and *HLA-C* haplotypes in a cohort of 3,496 individuals. Genetic makeup of Lithuania population is located within a European context, influenced by pre-Neolithic Western and Scandinavian hunting-gathering groups, early to middle Bronze Age steppe pastoralists, late Neolithic Bronze Age Europeans, and largely sheltered ^18^, which make it from an immune compatibility standpoint closely resembling European-American ^19^, and British populations ^20^. We compared this population to publicly available datasets of European ancestry and modeled the impact of gene editing on immune matching and population coverage, specifically in a pan-European context.

## 3. Materials and methods

### Ethical approval

This study is part of the ethical approval 2023/6-1524-984 “Highly-immune compatible iPS cells as source for Regenerative Medicine and Cell Therapy-oriented applications” from the Vilnius Regional Biomedical Research Ethics Committee to Vilnius University and 2023/4-1507-968 “Analysis of the Distribution of Human Leukocyte Antigen (HLA; Encoding Genes - HLA) Alleles and Haplotypes in the Group of the Lithuanian Unrelated Bone Marrow Donor Registry” to Vilnius University Hospital Santaros Klinikos. Written consents were obtained from the participants of the study.

### Genotyping

HLA typing for Registry donors’ peripheral blood was performed at the EFI accredited Immunogenetics laboratory at Vilnius University Hospital (Vilnius, Lithuania) using sequencing-based typing and at the ASHI accredited laboratory, HistoGenetics (Ossining, NY), using next-generation sequencing. Exons 2 and 3 for class I HLA were covered.

### Fibroblasts derivation and genotyping

Skin samples were collected using a 2-3 mm biopsy punch needle, and fragmented with a sterile scalpel and needle. Fibroblasts were grown with AmnioPrime Complete Medium (Capricorn Scientific cat no. APR-B), supplemented with Amphotericin B (Capricorn Scientific cat. no. AMP-B) for 21 to 45 days until fibroblast migrate from tissue sections and reach 80-90% confluence. The medium was regularly changed every 3 days to ensure optimal cell growth. Fibroblasts were routinely passaged with 0.25% Trypsin-EDTA at a density of 2 x 10^5^ cells/cm^2^. Genomic DNA from fibroblasts was purified using DNeasy blood and tissue kit (Qiagen cat no. 69504) and genotyped using the primers HLAA-P1: TCCAGGTGGACAGGTAAGGA, HLAA-P2: GTCACTGCCTGGGGTAGAAC, HLAB-P1: TGCATTCTGGGTTTCTCTACTGG, HLAB-P2: CACGCGAAACATCCCAATCA, HLAC-P1: AGGTAAGGCAAAGGGTGGGA, HLAC-P2: AGGCCGCCTGTACTTTTCTC. Samples were Sanger sequenced using the primers HLAA-P3: ACCCTCGTCCTGCTACTCTCG, HLAB-P3: ACCCTCCTCCTGCTGCTCTG, HLAC-P3: CGTTGGGGATTCTCCACTCC at Microsynth.

### Bioinformatics

Python and R scripts used for data analysis are made available thorough Supplementary Data and the open-source GitHub developer platform.

### Quantification of HLA allele frequency in the population

The total allele count in the dataset was divided by number of alleles (n=2) times the number of individuals in this study (n=3,496) having at least third-field resolution.

### Hardy-Weinberg Equilibrium (HWE) analyses

The observed genotypes present in the population were quantified (n=3,496). The allele frequencies were determined using sampled genotype count and the expected genotype frequencies calculated. The observed and expected genotype counts are compared with a *X*^2^ test. The *X*^2^ test is reliable for genotypes present more than 5 times in the population. Genotypes with count <5 times were filtered from the HWE analyses. The degrees of freedom df=(n(n+1)/2)−n are estimated in function of the number of possible genotypes and alleles number identified in the sampled population for each HLA class I gene, 44 for *HLA-A*, 83 for *HLA-B* and 45 for *HLA-C*.

### Regression analysis

Allele frequencies were extracted from the publicly available data from European-American ^19^, British ^20^ and African-American populations ^21^, and compared to the allele frequency from our study. Linear regression analyses y ∼ mx + c was performed using R for pairwise comparison of allele frequencies of *HLA-A*, *HLA-B* and *HLA-C*. Frequencies are calculated as frequency = allele count(dataset) / n(dataset).

### Principal component analysis

Monte Carlo population haplotypes were simulated based on the published allele frequencies of European-American, British and African-American cohort studies. Data was processed with one-hot encoding to convert allele entries per individual into 1 or 0, using the caret library from R ^22^. Centroids and Euclidean distances were calculated from the principal components. Distances were represented as edges and as heatmaps.

### HLA sequence analysis and sgRNA activity prediction

The sequences for all alleles in protein, transcript and gene level were downloaded as fasta files from IPD-IMGT-HLA database ^23^ and analyzed in python and R. Allele sequences were extracted based on the HLA alleles present in the population. Cas9 binding sites were extracted with python and analyzed in R using CrisprScore ^24^. Transmembrane prediction was conducted with DeepTMHMM ^25^.

## 4. Results

### Analysis of the HLA class I frequency in Lithuanian population

The Lithuanian Bone Marrow Donor Registry, located at Vilnius University Hospital Santaros Klinikos, includes 13,884 individuals, with 11,153 characterized at second field (protein level) for *HLA-A*, *HLA-B* and *HLA-C*. Of these, 3,496 individuals are characterized at third field (**Figure 1A**). We found that 858 individuals are at least homozygous for one HLA class I gene. A total of 542 individuals are homozygous for the coding sequence of *HLA-A*, 233 individuals are homozygous for *HLA-B*, and 338 individuals are homozygous for *HLA-C* (**Figure 1B**). We confirmed the Lithuanian population closely reassemble HLA distribution of the most frequent European HLA class I alleles (**Table 1** and **Figure 1**). The HLA types identified and their prevalence in the population are summarized in **Table 1**. The five most frequent *HLA-A* alleles are A*02:01:01, A*03:01:01, A*24:02:01, A*01:01:01 and A*11:01:01 which account for 74.6% of the population (**Figure 1C**). Noteworthy is the fact that HLA-A*02:01:01 is the most frequent HLA class I allele with a representation of 31.6% in the population. Similarly, the five most frequent *HLA-B* alleles are B*07:02:01, B*13:02:01, B*15:01:01, B*44:02:01 and B*40:01:01 which account for a 43.2% of the population (**Figure 1D**). HLA-B*07:02:01 alone represents 15.1% of the Lithuanian population. Furthermore, the five most frequent *HLA-C* alleles are C*07:02:01, C*06:02:01, C*04:01:01, C*02:02:02 and C*07:01:01 with a cumulative frequency of 59.2% of the population (**Figure 1E**).

**Figure 1.**
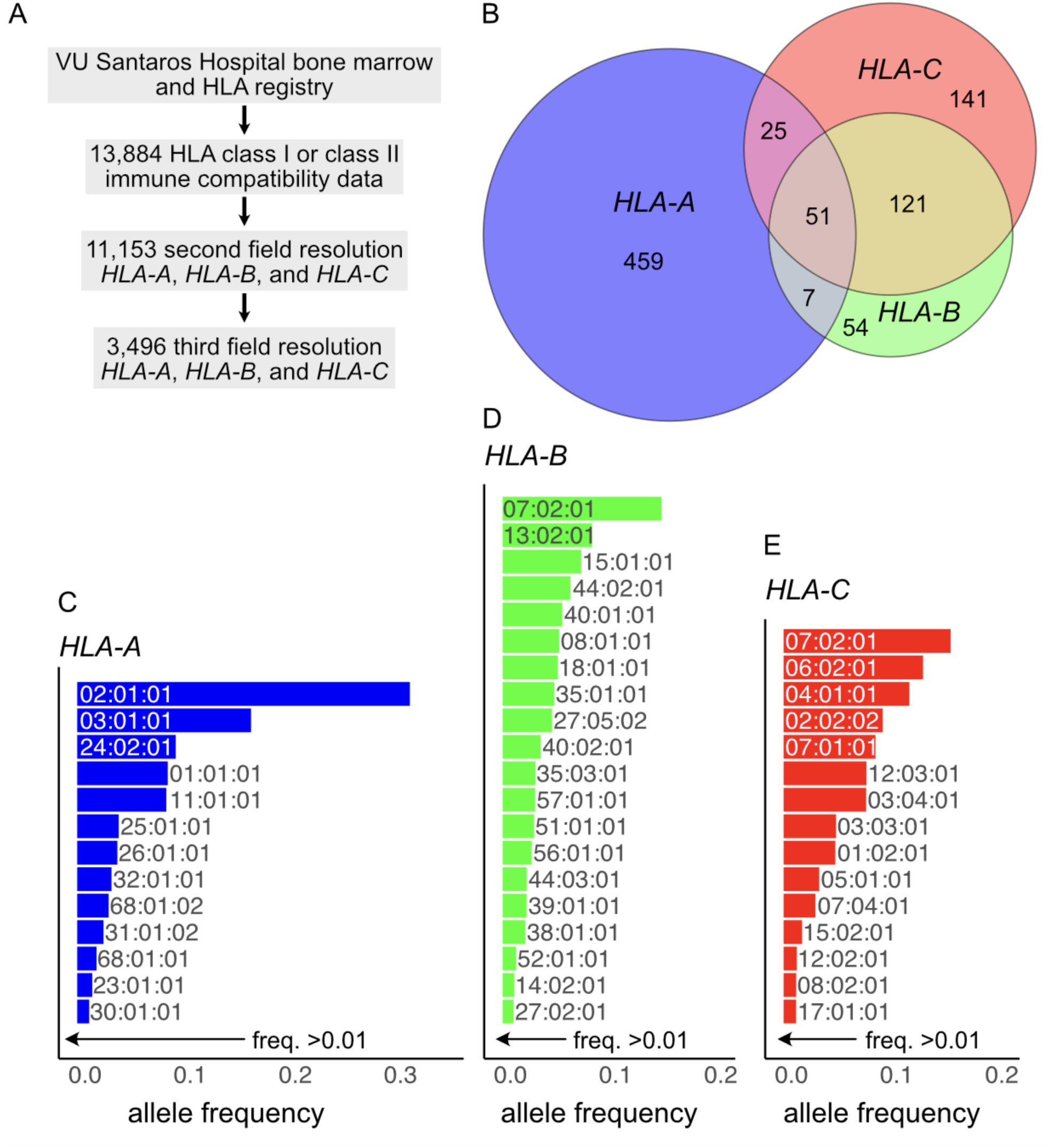
**A.** Dataset structure from this study **B.** Proportional Venn diagram showing the prevalence of HLA class I homozygous individuals in the Lithuanian population, with the composition of double homozygous and triple homozygous individuals highlighted. The most common HLA alleles with a frequency above 0.01, are shown for **C.** *HLA-A*, **D.** *HLA-B*, and **E.** *HLA-C*.

**Table 1.** HLA class I allele frequencies observed in the Lithuanian population (n = 3,496)

| HLA-A | count | freq | HLA-B | count | freq | HLA-C | count | freq |
| --- | --- | --- | --- | --- | --- | --- | --- | --- |
| 02:01:01 | 2212 | 0.3164 | 07:02:01 | 1057 | 0.1512 | 07:02:01 | 1110 | 0.1588 |
| 03:01:01 | 1156 | 0.1653 | 13:02:01 | 595 | 0.0851 | 06:02:01 | 927 | 0.1326 |
| 24:02:01 | 655 | 0.0937 | 15:01:01 | 523 | 0.0748 | 04:01:01 | 836 | 0.1196 |
| 01:01:01 | 602 | 0.0861 | 44:02:01 | 452 | 0.0646 | 02:02:02 | 659 | 0.0943 |
| 11:01:01 | 592 | 0.0847 | 40:01:01 | 397 | 0.0568 | 07:01:01 | 611 | 0.0874 |
| 25:01:01 | 276 | 0.0395 | 08:01:01 | 376 | 0.0538 | 12:03:01 | 551 | 0.0788 |
| 26:01:01 | 267 | 0.0382 | 18:01:01 | 368 | 0.0526 | 03:04:01 | 549 | 0.0785 |
| 32:01:01 | 228 | 0.0326 | 35:01:01 | 344 | 0.0492 | 03:03:01 | 349 | 0.0499 |
| 68:01:02 | 209 | 0.0299 | 27:05:02 | 329 | 0.0471 | 01:02:01 | 344 | 0.0492 |
| 31:01:02 | 175 | 0.0250 | 40:02:01 | 254 | 0.0363 | 05:01:01 | 237 | 0.0339 |
| 68:01:01 | 129 | 0.0184 | 35:03:01 | 219 | 0.0313 | 07:04:01 | 212 | 0.0303 |
| 23:01:01 | 100 | 0.0143 | 57:01:01 | 217 | 0.0310 | 15:02:01 | 123 | 0.0176 |
| 30:01:01 | 80 | 0.0114 | 51:01:01 | 209 | 0.0299 | 12:02:01 | 88 | 0.0126 |
| 29:02:01 | 64 | 0.0092 | 56:01:01 | 194 | 0.0277 | 08:02:01 | 84 | 0.0120 |
| 33:01:01 | 42 | 0.0060 | 44:03:01 | 161 | 0.0230 | 17:01:01 | 83 | 0.0119 |
| 66:01:01 | 29 | 0.0041 | 39:01:01 | 160 | 0.0229 | 14:02:01 | 56 | 0.0080 |
| 02:05:01 | 24 | 0.0034 | 38:01:01 | 151 | 0.0216 | 16:01:01 | 53 | 0.0076 |
| 68:02:01 | 24 | 0.0034 | 52:01:01 | 90 | 0.0129 | 03:02:01 | 21 | 0.0030 |
| 29:01:01 | 21 | 0.0030 | 14:02:01 | 78 | 0.0112 | 02:02:01 | 18 | 0.0026 |
| 33:03:01 | 21 | 0.0030 | 27:02:01 | 73 | 0.0104 | 16:02:01 | 12 | 0.0017 |
| 03:02:01 | 15 | 0.0021 | 49:01:01 | 65 | 0.0093 | 07:68 | 11 | 0.0016 |
| 02:06:01 | 9 | 0.0013 | 55:01:01 | 64 | 0.0092 | 15:05:01 | 10 | 0.0014 |
| 31:01:01 | 9 | 0.0013 | 37:01:01 | 63 | 0.0090 | 16:04:01 | 9 | 0.0013 |
| 30:04:01 | 7 | 0.0010 | 27:05:03 | 56 | 0.0080 | 07:02:08 | 4 | 0.0006 |
| 02:07:01 | 6 | 0.0009 | 41:02:01 | 56 | 0.0080 | 15:04:01 | 4 | 0.0006 |
| 02:35:01 | 6 | 0.0009 | 44:05:01 | 56 | 0.0080 | 08:01:01 | 3 | 0.0004 |
| 30:02:01 | 5 | 0.0007 | 35:02:01 | 52 | 0.0074 | 17:03:01 | 3 | 0.0004 |
| 03:22:01 | 4 | 0.0006 | 41:01:01 | 39 | 0.0056 | 18:01:01 | 3 | 0.0004 |
| 26:08:01 | 4 | 0.0006 | 51:07:01 | 31 | 0.0044 | 04:03:01 | 2 | 0.0003 |
| 31:08 | 3 | 0.0004 | 58:01:01 | 29 | 0.0041 | 04:04:01 | 2 | 0.0003 |
| 01:02:01 | 2 | 0.0003 | 50:01:01 | 26 | 0.0037 | 07:185 | 2 | 0.0003 |
| 02:30:01 | 2 | 0.0003 | 39:06:02 | 22 | 0.0031 | 12:98 | 2 | 0.0003 |
| 29:72 | 2 | 0.0003 | 35:08:01 | 19 | 0.0027 | 15:06:01 | 2 | 0.0003 |
| 69:01:01 | 2 | 0.0003 | 45:01:01 | 16 | 0.0023 | 03:54 | 1 | 0.0001 |
| 01:01:33 | 1 | 0.0001 | 47:01:01 | 13 | 0.0019 | 04:01:10 | 1 | 0.0001 |
| 02:02:01 | 1 | 0.0001 | 07:05:01 | 11 | 0.0016 | 05:16 | 1 | 0.0001 |
| 02:17:02 | 1 | 0.0001 | 18:03:01 | 10 | 0.0014 | 07:02:10 | 1 | 0.0001 |
| 02:38 | 1 | 0.0001 | 15:17:01 | 9 | 0.0013 | 07:75 | 1 | 0.0001 |
| 02:64:01 | 1 | 0.0001 | 15:24:01 | 9 | 0.0013 | 07:76:01 | 1 | 0.0001 |
| 03:01:14 | 1 | 0.0001 | 07:04:01 | 8 | 0.0011 | 08:03:01 | 1 | 0.0001 |
| 11:01:79 | 1 | 0.0001 | 57:03:01 | 7 | 0.0010 | 12:16:01 | 1 | 0.0001 |
| 24:03:01 | 1 | 0.0001 | 14:01:01 | 6 | 0.0009 | 15:09:01 | 1 | 0.0001 |
| 26:15 | 1 | 0.0001 | 44:27:01 | 5 | 0.0007 | 15:13:01 | 1 | 0.0001 |
| 33:05:01 | 1 | 0.0001 | 46:01:01 | 4 | 0.0006 | 16:15:01 | 1 | 0.0001 |
|  |  |  | 53:01:01 | 4 | 0.0006 | 16:201 | 1 | 0.0001 |
|  |  |  | 15:18:01 | 3 | 0.0004 |  |  |  |
|  |  |  | 15:39:01 | 3 | 0.0004 |  |  |  |
|  |  |  | 35:01:29 | 3 | 0.0004 |  |  |  |
|  |  |  | 39:06:01 | 3 | 0.0004 |  |  |  |
|  |  |  | 40:06:01 | 3 | 0.0004 |  |  |  |
|  |  |  | 48:01:01 | 3 | 0.0004 |  |  |  |
|  |  |  | 57:02:01 | 3 | 0.0004 |  |  |  |
|  |  |  | 07:07:01 | 2 | 0.0003 |  |  |  |
|  |  |  | 07:10:01 | 2 | 0.0003 |  |  |  |
|  |  |  | 13:01:01 | 2 | 0.0003 |  |  |  |
|  |  |  | 15:08:01 | 2 | 0.0003 |  |  |  |
|  |  |  | 15:16:01 | 2 | 0.0003 |  |  |  |
|  |  |  | 15:190:01 | 2 | 0.0003 |  |  |  |
|  |  |  | 39:24:01 | 2 | 0.0003 |  |  |  |
|  |  |  | 40:01:02 | 2 | 0.0003 |  |  |  |
|  |  |  | 40:12:01 | 2 | 0.0003 |  |  |  |
|  |  |  | 40:94 | 2 | 0.0003 |  |  |  |
|  |  |  | 44:03:02 | 2 | 0.0003 |  |  |  |
|  |  |  | 44:48:01 | 2 | 0.0003 |  |  |  |
|  |  |  | 51:08:01 | 2 | 0.0003 |  |  |  |
|  |  |  | 08:262 | 1 | 0.0001 |  |  |  |
|  |  |  | 15:01:10 | 1 | 0.0001 |  |  |  |
|  |  |  | 15:09:01 | 1 | 0.0001 |  |  |  |
|  |  |  | 15:34:01 | 1 | 0.0001 |  |  |  |
|  |  |  | 15:73:01 | 1 | 0.0001 |  |  |  |
|  |  |  | 27:03 | 1 | 0.0001 |  |  |  |
|  |  |  | 27:07:01 | 1 | 0.0001 |  |  |  |
|  |  |  | 27:12:01 | 1 | 0.0001 |  |  |  |
|  |  |  | 39:68 | 1 | 0.0001 |  |  |  |
|  |  |  | 40:01:20 | 1 | 0.0001 |  |  |  |
|  |  |  | 40:32 | 1 | 0.0001 |  |  |  |
|  |  |  | 42:01:01 | 1 | 0.0001 |  |  |  |
|  |  |  | 42:02:01 | 1 | 0.0001 |  |  |  |
|  |  |  | 44:04 | 1 | 0.0001 |  |  |  |
|  |  |  | 47:02 | 1 | 0.0001 |  |  |  |
|  |  |  | 51:01:19 | 1 | 0.0001 |  |  |  |
|  |  |  | 51:05:01 | 1 | 0.0001 |  |  |  |
|  |  |  | 52:01:02 | 1 | 0.0001 |  |  |  |

It is important to highlight that the *HLA-B* gene exhibits the largest diversity of alleles, followed by *HLA-A* and *HLA-C* (**Table 1**), as also observed in previous studies on populations of European origin ^19,20^. Comparisons of the Lithuanian Class I HLA frequencies with those reported for the European-American and British populations through linear regression models show strong correlations between the three cohorts (**Figure 2**). The linear regression analyses show an average slope of 0.914 for *HLA-A*, 0.826 for *HLA-B*, and 0.860 for *HLA-C*. This indicates the populations closely resemble each other, regarding the composition and prevalence of allele variants. In order to validate this comparison we fitted a linear model with the HLA class I frequencies of African-American population ^21^ and confirmed the allele frequency differences as reported by regressions, with slope of 0.346 for *HLA-A*, 0.310 for *HLA-B*, and 0.611 for *HLA-C* (**Figure 2**). Principal component analysis (PCA) of the genotypes of the Lithuanian population and those extracted from published datasets, reconstructed through Monte Carlo analysis based on reported allele frequencies, showed close proximity (**Figure 3A**). The Euclidean distances between the centroids of the populations were quantified and represented in the PCA and as a heatmap (**Figure 3B**). The distance metrics indicate that the centroid of the Lithuanian population is proximal to the European-American and British populations with distances of 2.49 and 2.95 relative units, respectively. The British and European-American population closely resemble each other with a Euclidean distance of 0.61. Conversely, the African-American population presents an average Euclidean distance of 5.72 relative units with the other datasets.

**Figure 2.**
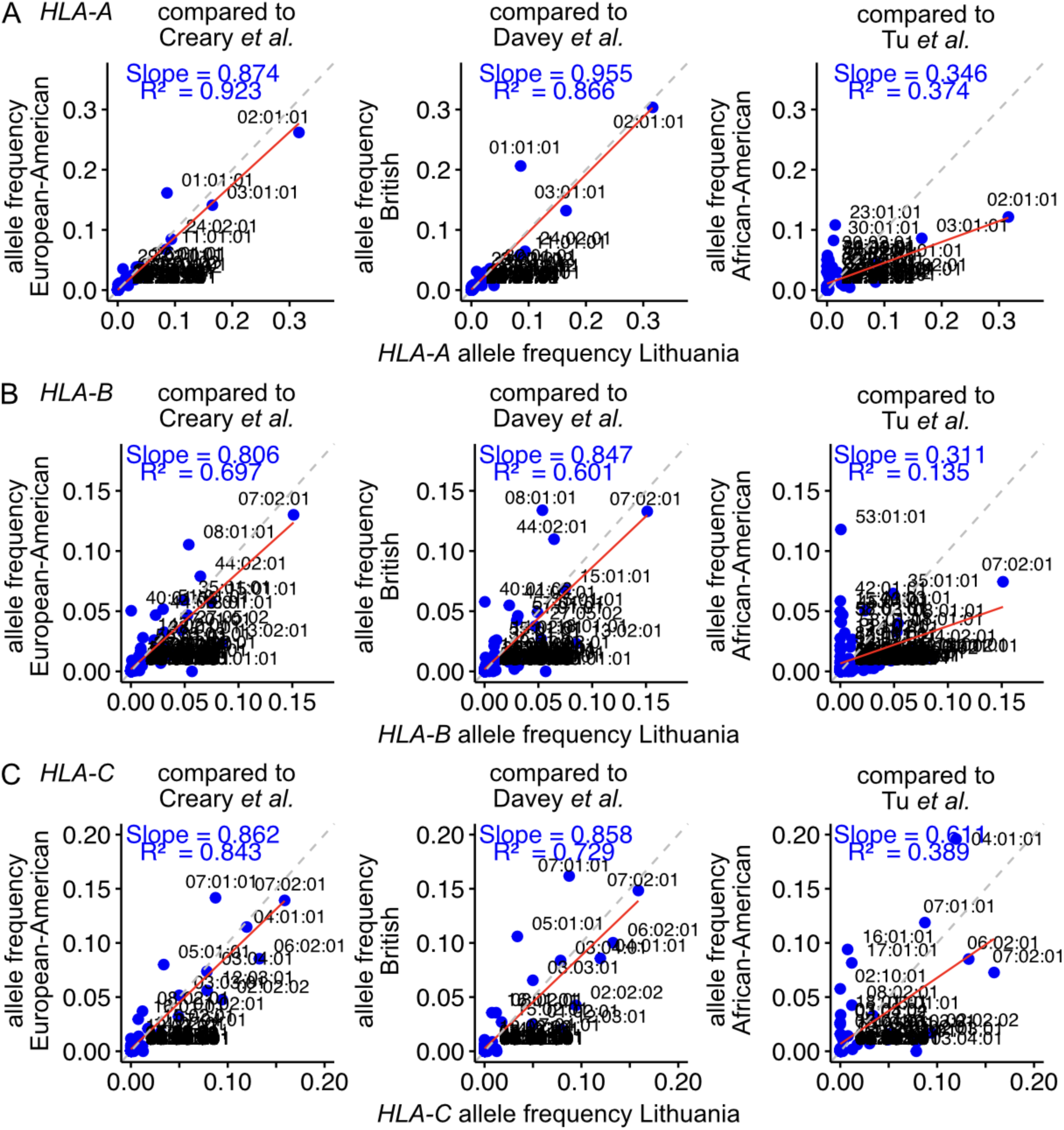
Comparison of the HLA allele frequencies identified in the Lithuanian population with those reported in studies for the European-American population, the British population and the African-American population for **A.** the *HLA-A* transcript, **B.** *HLA-B* transcript and **C.** the *HLA-C* transcript. Reference lines with slope n=1 are represented in dashed grey. The linear regressions of frequencies on the scatter plots are represented with a red solid line, with the R^2^ of the linear model and the slope indicated.

**Figure 3.**
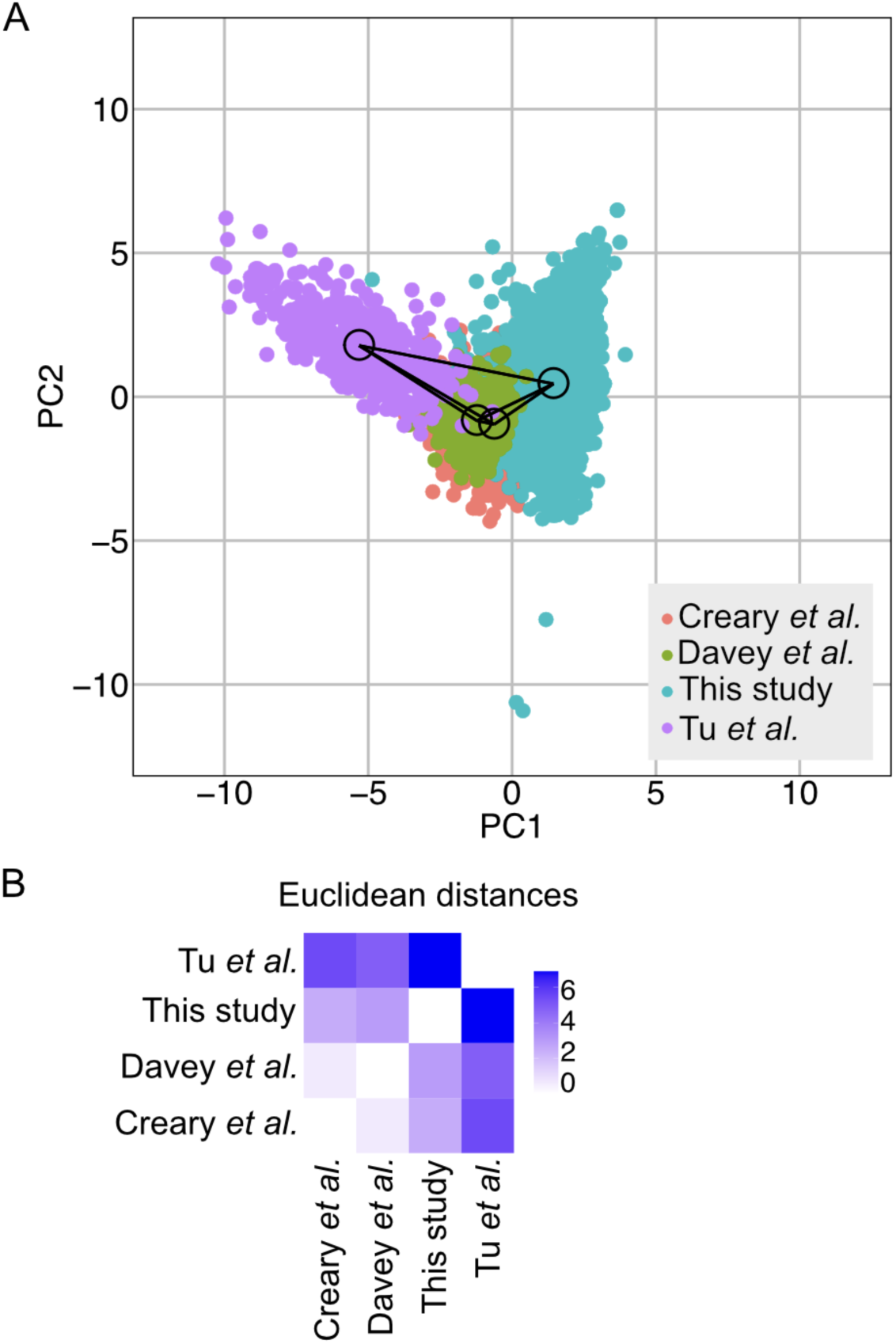
**A.** Principal component analysis of the HLA class I distribution in the Lithuanian population (this study) and pan-European populations, including European-American, British, and African-American cohorts. The centroid of each population is marked with a circle. The Euclidean distances between the centroids were calculated, and their edges are plotted with solid lines. **B.** Euclidean distance heatmap between the studied populations. Blue correspond to higher Euclidean distances in the principal component space.

### Population analysis for HLA class I composition

We conducted a Hardy-Weinberg equilibrium analysis to determine whether there were deviations in the genotype frequencies of the Lithuanian HLA class I allele pool compared to the expected frequencies under Hardy-Weinberg conditions for each allele. We found 44 alleles for *HLA-A*, 83 alleles for *HLA-B*, and 45 alleles for *HLA-C* within the 3,496 individuals analyzed (**Table 1**). This diversity of allele types results in a maximum of 990 genotypes for *HLA-A*, 3,486 for *HLA-B*, and 1,035 for *HLA-C*. When excluding the rare variants, defined as those with a frequency of less than 0.01 in the population, we found that 22 genotypes deviate from Hardy-Weinberg equilibrium for *HLA-A*, 16 deviate for *HLA-B*, and 30 deviate for *HLA-C* (**Figure 4A**). The highest-ranked *HLA-A* genotype that is present more frequently than expected is HLA-A*03:01:01-25:01:01, which combines two of the top six most abundant allele types (**Figure 4D**). Likewise, HLA-B*13:02:01-27:05:02, HLA-B*08:01:01-15:01:01, and HLA-B*15:01:01-44:02:01, which are combinations of the top ten most frequent allele types, are present at higher frequencies than expected under Hardy-Weinberg equilibrium conditions (**Figure 4A**). Furthermore, a larger group of *HLA-C* genotypes deviate from equilibrium, including genotypes of the most frequent allele types HLA-C*07:01:01-12:03:01, HLA-C*01:02:01-04:01:01, HLA-C*02:02:02-06:02:01, HLA-C*04:01:01-06:02:01, HLA-C*03:04:01-12:03:01, HLA-C*01:02:01-06:02:01, and HLA-C*07:02:01-07:02:01 (**Figure 4A**). In fact, HLA-C*07:02:01 is the most frequent allele type, and its homozygous combination is enriched more than expected based on Hardy-Weinberg equilibrium conditions.

**Figure 4.**
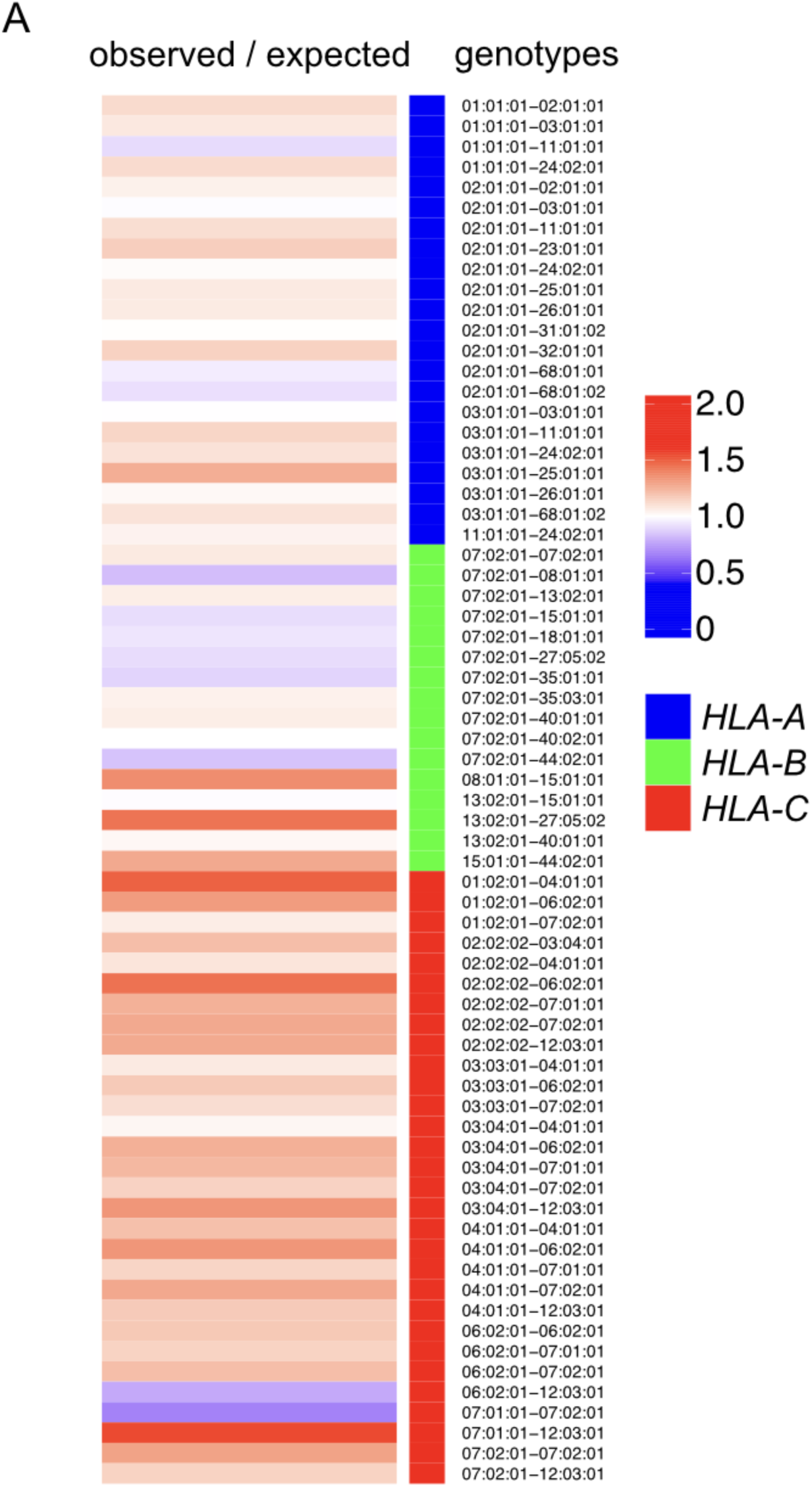
Hardy-Weinberg equilibrium (HWE) analysis for **A.** *HLA-A*, *HLA-B* and *HLA-C* genotypes. The heatmaps represent the ratio between observed genotype frequencies and expected genotype frequencies. Only the genotypes were *X*^2^ value exceeds the *X*^2^-threshold, indicating a HWE deviation, were filtered. The subset of genotypes with a frequency higher than 0.01 are represented in heatmaps.

### Identification of HLA class I double homozygous and triple homozygous individuals in the Lithuanian population

Of the HLA homozygous individuals, a total of 153 are double homozygous (**Figure 1A** and **Table 2**), 58 are double homozygous for *HLA-A* and *HLA-B*, 76 are double homozygous for *HLA-A* and *HLA-C*, and 172 are double homozygous for *HLA-B* and *HLA-C*. Remarkably, 51 individuals are triple homozygous for *HLA-A*, *HLA-B*, and *HLA-C* (**Figure 1A** and **Table 3**). Haplotype frequencies of the complete dataset (3,496 individuals) are available in Supplementary Data.

**Table 2.** HLA class I double homozygous haplotypes identified in this study (n = 153)

| haplotype | count |
| --- | --- |
| A*02:01:01 / 03:01:01 - B*07:02:01 / 07:02:01 - C*07:02:01 / 07:02:01 | 29 |
| A*03:01:01 / 11:01:01 - B*07:02:01 / 07:02:01 - C*07:02:01 / 07:02:01 | 10 |
| A*03:01:01 / 24:02:01 - B*07:02:01 / 07:02:01 - C*07:02:01 / 07:02:01 | 8 |
| A*02:01:01 / 02:01:01 - B*13:02:01 / 57:01:01 - C*06:02:01 / 06:02:01 | 6 |
| A*02:01:01 / 11:01:01 - B*07:02:01 / 07:02:01 - C*07:02:01 / 07:02:01 | 4 |
| A*03:01:01 / 11:01:01 - B*35:01:01 / 35:01:01 - C*04:01:01 / 04:01:01 | 3 |
| A*01:01:01 / 02:01:01 - B*13:02:01 / 13:02:01 - C*06:02:01 / 06:02:01 | 2 |
| A*02:01:01 / 02:01:01 - B*15:01:01 / 35:03:01 - C*04:01:01 / 04:01:01 | 2 |
| A*02:01:01 / 02:01:01 - B*15:01:01 / 40:01:01 - C*03:04:01 / 03:04:01 | 2 |
| A*02:01:01 / 03:01:01 - B*13:02:01 / 13:02:01 - C*06:02:01 / 06:02:01 | 2 |
| A*02:01:01 / 03:01:01 - B*40:01:01 / 40:01:01 - C*03:04:01 / 03:04:01 | 2 |
| A*02:01:01 / 11:01:01 - B*15:01:01 / 15:01:01 - C*04:01:01 / 04:01:01 | 2 |
| A*02:01:01 / 24:02:01 - B*13:02:01 / 13:02:01 - C*06:02:01 / 06:02:01 | 2 |
| A*02:01:01 / 25:01:01 - B*08:01:01 / 08:01:01 - C*07:01:01 / 07:01:01 | 2 |
| A*02:01:01 / 26:01:01 - B*07:02:01 / 07:02:01 - C*07:02:01 / 07:02:01 | 2 |
| A*02:01:01 / 30:01:01 - B*13:02:01 / 13:02:01 - C*06:02:01 / 06:02:01 | 2 |
| A*03:01:01 / 26:01:01 - B*35:01:01 / 35:01:01 - C*04:01:01 / 04:01:01 | 2 |
| A*03:01:01 / 31:01:02 - B*39:01:01 / 39:01:01 - C*12:03:01 / 12:03:01 | 2 |
| A*03:01:01 / 32:01:01 - B*07:02:01 / 07:02:01 - C*07:02:01 / 07:02:01 | 2 |
| A*11:01:01 / 26:01:01 - B*07:02:01 / 07:02:01 - C*07:02:01 / 07:02:01 | 2 |
| A*01:01:01 / 02:01:01 - B*08:01:01 / 08:01:01 - C*07:01:01 / 07:01:01 | 1 |
| A*01:01:01 / 02:01:01 - B*35:03:01 / 35:03:01 - C*04:01:01 / 04:01:01 | 1 |
| A*01:01:01 / 02:01:01 - B*40:01:01 / 40:01:01 - C*03:04:01 / 03:04:01 | 1 |
| A*01:01:01 / 02:01:01 - B*57:01:01 / 57:01:01 - C*06:02:01 / 06:02:01 | 1 |
| A*01:01:01 / 23:01:01 - B*44:03:01 / 44:03:01 - C*04:01:01 / 04:01:01 | 1 |
| A*01:01:01 / 24:02:01 - B*07:02:01 / 07:02:01 - C*07:02:01 / 07:02:01 | 1 |
| A*01:01:01 / 24:02:01 - B*13:02:01 / 13:02:01 - C*06:02:01 / 06:02:01 | 1 |
| A*02:01:01 / 02:01:01 - B*07:02:01 / 15:01:01 - C*03:04:01 / 03:04:01 | 1 |
| A*02:01:01 / 02:01:01 - B*07:02:01 / 39:01:01 - C*07:02:01 / 07:02:01 | 1 |
| A*02:01:01 / 02:01:01 - B*08:01:01 / 18:01:01 - C*07:01:01 / 07:01:01 | 1 |
| A*02:01:01 / 02:01:01 - B*15:01:01 / 15:01:01 - C*03:03:01 / 04:01:01 | 1 |
| A*02:01:01 / 02:01:01 - B*15:01:01 / 15:01:01 - C*03:04:01 / 04:01:01 | 1 |
| A*02:01:01 / 02:01:01 - B*27:05:02 / 27:05:02 - C*01:02:01 / 02:02:02 | 1 |
| A*02:01:01 / 02:01:01 - B*27:05:02 / 51:01:01 - C*01:02:01 / 01:02:01 | 1 |
| A*02:01:01 / 02:01:01 - B*27:05:02 / 56:01:01 - C*01:02:01 / 01:02:01 | 1 |
| A*02:01:01 / 02:01:01 - B*38:01:01 / 39:01:01 - C*12:03:01 / 12:03:01 | 1 |
| A*02:01:01 / 02:01:01 - B*44:02:01 / 44:02:01 - C*03:03:01 / 07:04:01 | 1 |
| A*02:01:01 / 02:01:01 - B*44:02:01 / 44:02:01 - C*05:01:01 / 07:04:01 | 1 |
| A*02:01:01 / 02:01:01 - B*44:05:01 / 57:01:01 - C*02:02:02 / 02:02:02 | 1 |
| A*02:01:01 / 02:01:01 - B*51:01:01 / 51:01:01 - C*02:02:02 / 15:02:01 | 1 |
| A*02:01:01 / 03:01:01 - B*15:01:01 / 15:01:01 - C*03:03:01 / 03:03:01 | 1 |
| A*02:01:01 / 03:01:01 - B*18:01:01 / 18:01:01 - C*12:03:01 / 12:03:01 | 1 |
| A*02:01:01 / 03:01:01 - B*44:02:01 / 44:02:01 - C*07:04:01 / 07:04:01 | 1 |
| A*02:01:01 / 03:01:01 - B*56:01:01 / 56:01:01 - C*01:02:01 / 01:02:01 | 1 |
| A*02:01:01 / 11:01:01 - B*40:02:01 / 40:02:01 - C*02:02:01 / 02:02:01 | 1 |
| A*02:01:01 / 24:02:01 - B*07:02:01 / 07:02:01 - C*07:02:01 / 07:02:01 | 1 |
| A*02:01:01 / 24:02:01 - B*15:01:01 / 15:01:01 - C*03:03:01 / 03:03:01 | 1 |
| A*02:01:01 / 26:01:01 - B*13:02:01 / 13:02:01 - C*06:02:01 / 06:02:01 | 1 |
| A*02:01:01 / 26:01:01 - B*15:01:01 / 15:01:01 - C*03:03:01 / 03:03:01 | 1 |
| A*02:01:01 / 26:01:01 - B*27:05:02 / 27:05:02 - C*01:02:01 / 01:02:01 | 1 |
| A*02:01:01 / 26:01:01 - B*38:01:01 / 38:01:01 - C*12:03:01 / 12:03:01 | 1 |
| A*02:01:01 / 26:01:01 - B*40:01:01 / 40:01:01 - C*03:04:01 / 03:04:01 | 1 |
| A*02:01:01 / 31:01:02 - B*13:02:01 / 13:02:01 - C*06:02:01 / 06:02:01 | 1 |
| A*02:01:01 / 31:01:02 - B*40:01:01 / 40:01:01 - C*03:04:01 / 03:04:01 | 1 |
| A*02:01:01 / 32:01:01 - B*40:02:01 / 40:02:01 - C*02:02:02 / 02:02:02 | 1 |
| A*02:01:01 / 68:01:01 - B*40:01:01 / 40:01:01 - C*03:04:01 / 03:04:01 | 1 |
| A*02:01:01 / 68:01:02 - B*15:01:01 / 15:01:01 - C*03:04:01 / 03:04:01 | 1 |
| A*02:01:01 / 68:01:02 - B*35:03:01 / 35:03:01 - C*04:01:01 / 04:01:01 | 1 |
| A*02:01:01 / 68:01:02 - B*40:01:01 / 40:01:01 - C*03:04:01 / 03:04:01 | 1 |
| A*03:01:01 / 03:01:01 - B*07:02:01 / 07:02:01 - C*07:02:01 / 07:185:01 | 1 |
| A*03:01:01 / 03:01:01 - B*07:02:01 / 07:04:01 - C*07:02:01 / 07:02:01 | 1 |
| A*03:01:01 / 03:01:01 - B*08:01:01 / 47:01:01 - C*07:01:01 / 07:01:01 | 1 |
| A*03:01:01 / 03:01:01 - B*18:01:01 / 35:03:01 - C*12:03:01 / 12:03:01 | 1 |
| A*03:01:01 / 11:01:01 - B*56:01:01 / 56:01:01 - C*01:02:01 / 01:02:01 | 1 |
| A*03:01:01 / 24:02:01 - B*35:01:01 / 35:01:01 - C*04:01:01 / 04:01:01 | 1 |
| A*03:01:01 / 24:02:01 - B*40:02:01 / 40:02:01 - C*02:02:02 / 02:02:02 | 1 |
| A*03:01:01 / 25:01:01 - B*35:01:01 / 35:01:01 - C*04:01:01 / 04:01:01 | 1 |
| A*03:01:01 / 26:01:01 - B*40:01:01 / 40:01:01 - C*03:04:01 / 03:04:01 | 1 |
| A*03:01:01 / 30:01:01 - B*07:02:01 / 07:02:01 - C*07:02:01 / 07:02:01 | 1 |
| A*03:01:01 / 68:01:02 - B*07:02:01 / 07:02:01 - C*07:02:01 / 07:02:01 | 1 |
| A*03:01:14 / 31:01:02 - B*07:02:01 / 07:02:01 - C*07:02:01 / 07:02:01 | 1 |
| A*11:01:01 / 11:01:01 - B*15:01:01 / 51:01:01 - C*04:01:01 / 04:01:01 | 1 |
| A*11:01:01 / 11:01:01 - B*35:01:01 / 44:03:01 - C*04:01:01 / 04:01:01 | 1 |
| A*11:01:01 / 24:02:01 - B*07:02:01 / 07:02:01 - C*07:02:01 / 07:02:01 | 1 |
| A*11:01:01 / 25:01:01 - B*56:01:01 / 56:01:01 - C*01:02:01 / 01:02:01 | 1 |
| A*11:01:01 / 26:01:01 - B*40:02:01 / 40:02:01 - C*02:02:02 / 02:02:02 | 1 |
| A*11:01:01 / 31:01:02 - B*51:01:01 / 51:01:01 - C*15:02:01 / 15:02:01 | 1 |
| A*11:01:01 / 68:01:02 - B*07:02:01 / 07:02:01 - C*07:02:01 / 07:02:01 | 1 |
| A*24:02:01 / 24:02:01 - B*07:02:01 / 39:06:02 - C*07:02:01 / 07:02:01 | 1 |
| A*24:02:01 / 24:02:01 - B*08:01:01 / 39:01:01 - C*07:02:01 / 07:02:01 | 1 |
| A*24:02:01 / 24:02:01 - B*13:02:01 / 37:01:01 - C*06:02:01 / 06:02:01 | 1 |
| A*24:02:01 / 25:01:01 - B*18:01:01 / 18:01:01 - C*12:03:01 / 12:03:01 | 1 |
| A*25:01:01 / 30:01:01 - B*13:02:01 / 13:02:01 - C*06:02:01 / 06:02:01 | 1 |
| A*25:01:01 / 31:01:02 - B*40:01:01 / 40:01:01 - C*03:04:01 / 03:04:01 | 1 |
| A*26:01:01 / 32:01:01 - B*38:01:01 / 38:01:01 - C*12:03:01 / 12:03:01 | 1 |

**Table 3.** HLA class I triple homozygous haplotypes identified in this study (n = 51)

| Haplotype HLA-A, HLA-B, and HLA-C triple homozygous | count |
| --- | --- |
| A*03:01:01 - B*07:02:01 - C*07:02:01 | 15 |
| A*02:01:01 - B*13:02:01 - C*06:02:01 | 10 |
| A*01:01:01 - B*08:01:01 - C*07:01:01 | 6 |
| A*02:01:01 - B*07:02:01 - C*07:02:01 | 4 |
| A*02:01:01 - B*40:01:01 - C*03:04:01 | 3 |
| A*02:01:01 - B*57:01:01 - C*06:02:01 | 2 |
| A*03:01:01 - B*56:01:01 - C*01:02:01 | 2 |
| A*25:01:01 - B*18:01:01 - C*12:03:01 | 2 |
| A*02:01:01 - B*15:01:01 - C*03:04:01 | 1 |
| A*02:01:01 - B*27:05:02 - C*02:02:02 | 1 |
| A*25:01:01 - B*35:01:01 - C*04:01:01 | 1 |
| A*26:01:01 - B*38:01:01 - C*12:03:01 | 1 |
| A*31:01:02 - B*51:01:01 - C*05:01:01 | 1 |
| A*68:01:01 - B*40:01:01 - C*03:04:01 | 1 |
| A*68:01:02 - B*44:02:01 - C*07:04:01 | 1 |

### HLA class I immune compatibility cumulative-coverage based on stochastic sampling, double homozygous or triple homozygous sampling

The cumulative-coverage accounts for the extent of redundancy in the population, which allows for matching even when a single sample may not be accessible for donation. Using the third-field HLA information for *HLA-A*, *HLA-B* and *HLA-C* from 3,496 individuals from this study, we conducted sampling simulations and calculated the cumulative coverage of 1,000 randomly selected individuals from the Lithuanian population. Using 100 simulations, we found that, on average, 1,000 randomly selected samples achieve a cumulative-coverage of 3.1 ± 0.4 times the population (**Figure 5A**). We found that approximately 329 individuals provide an average cumulative-coverage of 0.99 ± 0.2 of the populations (**Figure 5B**). We then excluded the possibility of autologous donation from the population’s cumulative-coverage analysis. When randomly selecting 1,000 individuals in 100 sampling iterations, a lower cumulative-coverage was achieved, with an average of 2.7 ± 0.4 times the population (**Figure 5C**). Sampling 329 individuals, excluding the possibility of autologous donation, results in a cumulative-coverage of only 0.9 ± 0.2 times the population (**Figure 5D**). Remarkably, when evaluating the cumulative-coverage of double or triple homozygous for *HLA-A*, *HLA-B* and *HLA-C*, a higher cumulative-coverage is achieved with a smaller set of samples. The 153 double homozygous samples achieve a cumulative-coverage of 2.2 times the population, and the 51 triple homozygous samples yield a cumulative-coverage of 4.9 times the population (**Figure 5E**). A side-by-side comparison of the double and triple homozygous cumulative-coverage with stochastic sampling of 1,000 individuals highlights the impact of homozygosity on immune compatibility to the population (**Figure 5E**).

**Figure 5.**
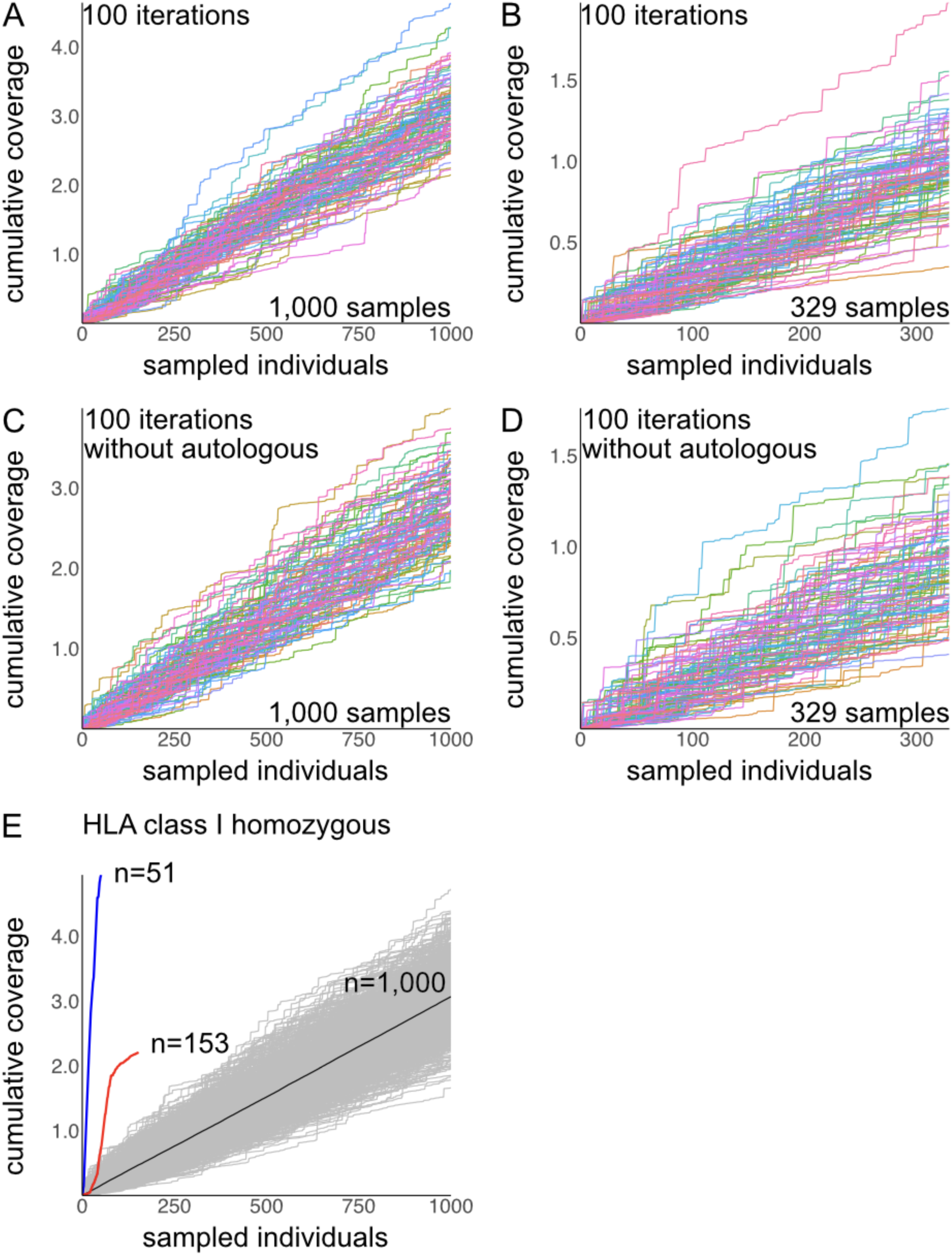
Cumulative-coverage of HLA class I immune matching in the Lithuanian population. Individuals in our dataset were sampled, and their cumulative-coverage in the population was calculated with 100 sampling iterations for **A.** 1,000 individuals, **B.** 329 individuals, which is the estimated sample size to reach a 1-time cumulative-coverage. **C.** Sampling of 1,000 and **D.** 329 individuals excluding the possibility of autologous donation. **E.** Cumulative-coverage for the individuals found to be double homozygous (red) and triple homozygous (blue) for *HLA-A*, *HLA-B* and *HLA-C*. Comparison with 1,000 randomly sampled individuals in 100 iterations (grey). The average cumulative coverage of all iterations is shown in black.

### Compatibility of HLA class I in the Lithuanian and pan-European populations

We stochastically arranged the 3,496 donors and interrogated whether the subset of *HLA-A*, *HLA-B* and *HLA-C* triple homozygous (51 samples), and double homozygous (153 samples) were compatible with the 3,496 patients (**Figure 6A**). We found that our cohort of triple homozygous patients matches 60.46% of the Lithuanian population. Likewise, the double homozygous cohort matches 33.32% of the Lithuanian population. In comparison, a randomly selected subset of 153 or 51 samples of the dataset could match only 11.84% (**Figure 6B**) and 4.1% (**Figure 6C**) of the Lithuanian population, respectively. We then evaluated the matching provided by our triple homozygous and double homozygous cohorts to the pan-European population by assessing their immune compatibility with Monte Carlo datasets, reconstructed from the allele frequencies reported for European-American and British individuals. Remarkably, we found that the 51 triple homozygous samples of our cohort match 13.4% of the British population, while the double homozygous cohort matches 5.2% (**Figure 6D**). Additionally, we found that triple homozygous samples match 7.4% of the European-American population, and double homozygous samples match of 3.3% (**Figure 6E**).

**Figure 6.**
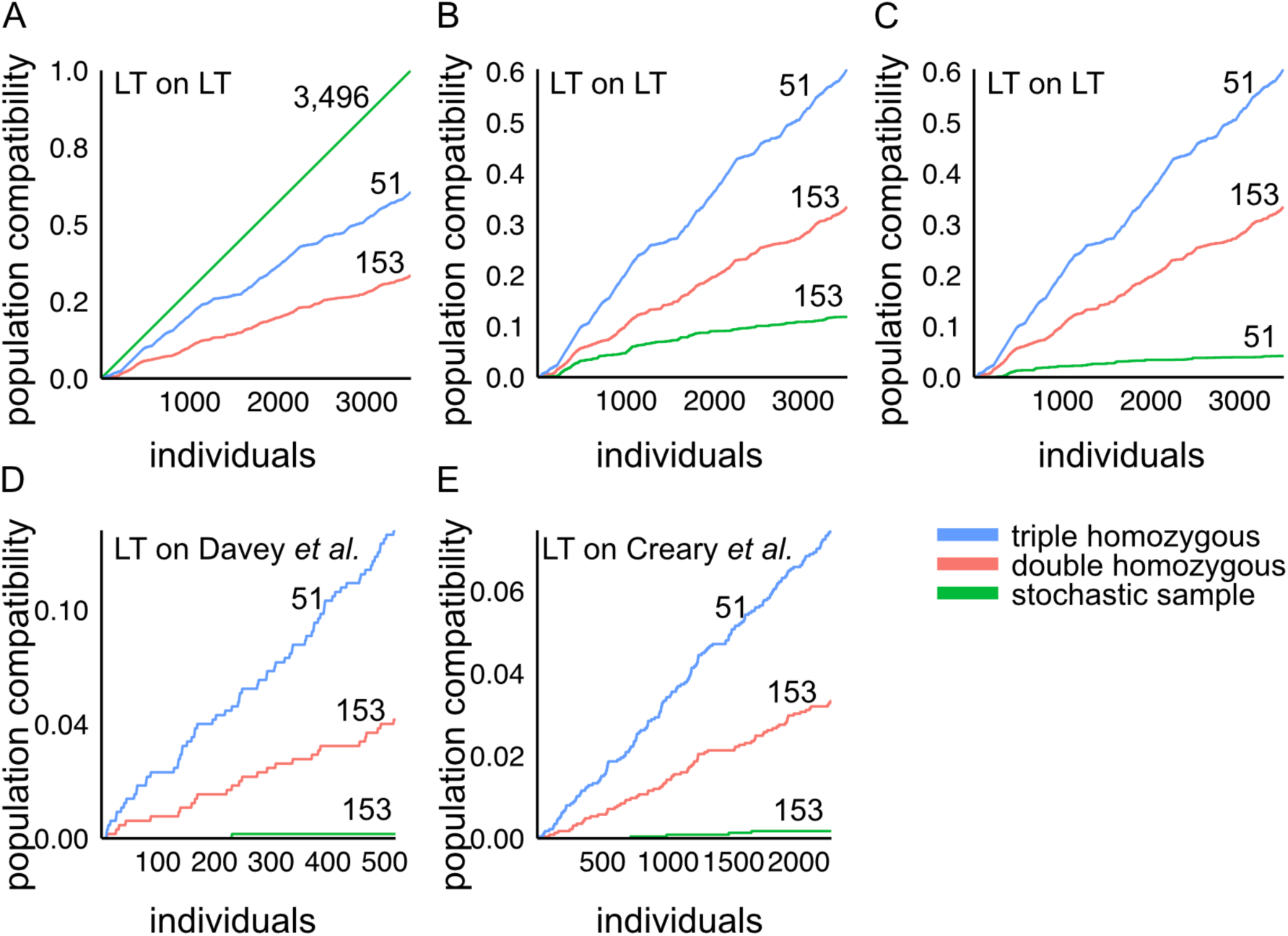
Population compatibility of *HLA-A*, *HLA-B* and *HLA-C* genotypes in Lithuanian samples with the Lithuanian and pan-European populations. Immune compatibility of triple homozygous (51 individuals), double homozygous (153 individuals) and **A.** all samples from the cohort of 3,496 individuals of this study, **B.** stochastically selected samples from 153 individuals, and **C.** stochastically selected samples from 51 individuals. Immune compatibility of HLA class I genes, *HLA-A*, *HLA-B* and *HLA-C*, in Lithuanian samples with **D.** British datasets and **E.** European-American datasets. The triple homozygous individuals are indicated in blue, double homozygous individuals in red, and stochastically selected subsamples in green.

### Cas9 activity prediction on HLA class I alleles in the Lithuanian population

We extracted the Cas9 binding site sequences from the HLA alleles present in the Lithuanian population. First, we focused on the analysis of target regions encompassing the gene body from 5’UTR to the 3’UTR. We found 1,996 unique target sites in *HLA-A*, 2,342 unique target sites in *HLA-B*, and 2,300 unique target sites in *HLA-C*. We calculated the activity prediction score based on the rule set 1 of nuclease catalytic activity ^26^. We found that, as in non-hyper polymorphic genes, the activity scores of all HLA alleles are centered in the non-active Q4 quadrant. We show this distribution for the five most frequent alleles of *HLA-A*, *HLA-B* and *HLA-C* (**Figure 7A**). The potential of HLA gene knock-out to modulate immune compatibility is well accepted in the literature. Although pairs of guide RNAs can be used in conjunction to create exon spanning knock-outs, we focused on guide RNA in exon regions. From the guide RNAs present in the gene body, we found 679 unique target sites in the *HLA-A* exons of Lithuanian alleles, 698 in *HLA-B*, and 687 in *HLA-C* (**Figure 7B**). Since *HLA-A*, *HLA-B* and *HLA-C* are class I single-span transmembrane proteins (**Figure 7C**), only guide RNAs targeting the ectodomain have the capacity to create knock-outs that eliminate plasma membrane expression of HLA genes. We predicted the transmembrane spanning region ^25^ of the allele sequences and focused on guide RNAs directed to the N-terminus, upstream of the predicted transmembrane domain. We found there are 615 unique target sites in Lithuanian alleles on *HLA-A* ectodomains, 658 on *HLA-B* and 613 on *HLA-C* (**Figure 7B**). Of those useful for ectodomain targeting, a fraction have predicted high activity score larger than 0.5. These include 54 for *HLA-A*, 75 for *HLA-B,* and 66 for *HLA-C* (**Figure 7B**).

**Figure 7.**
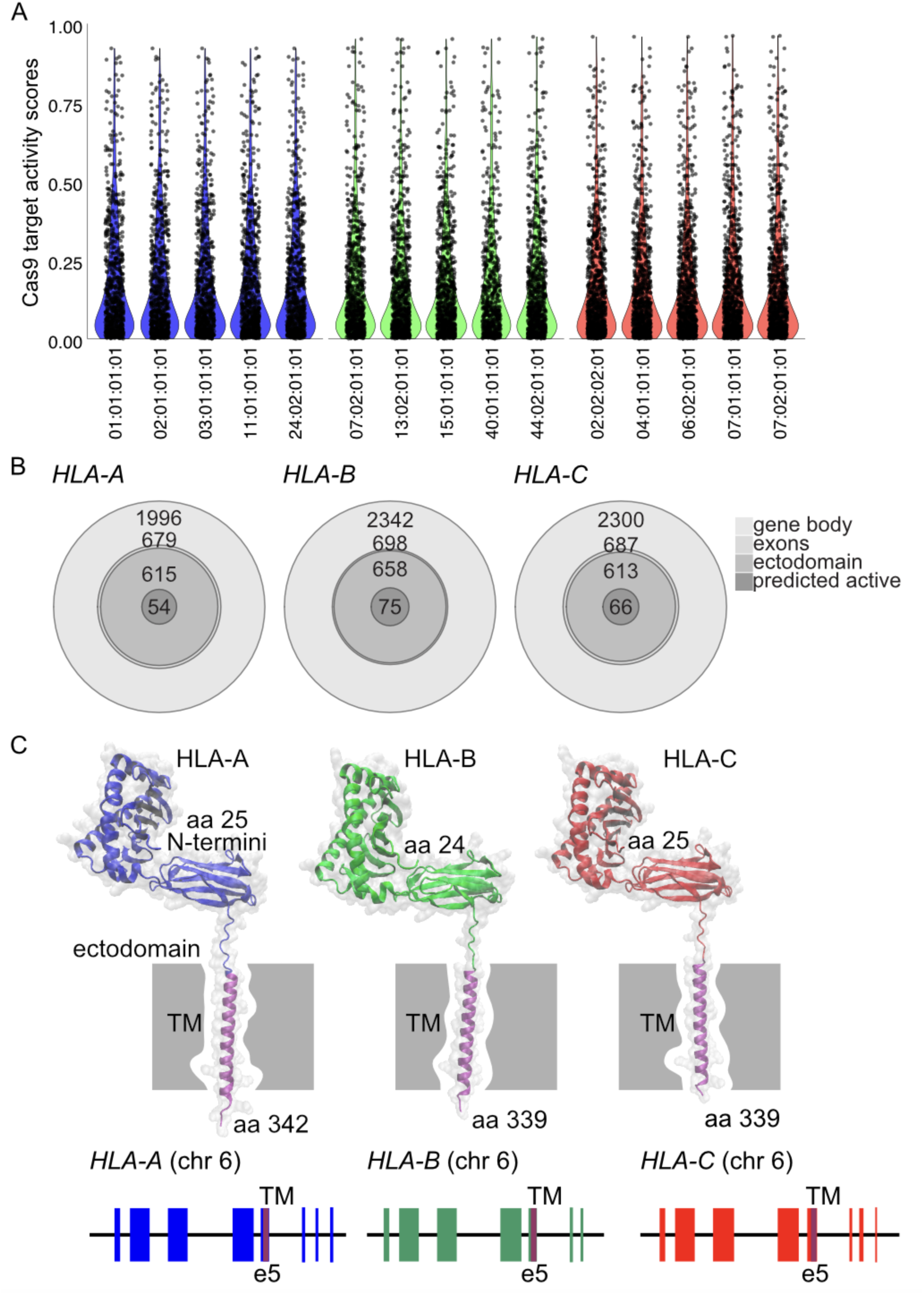
Predicted guide RNA sequence activity for the 5 most frequent alleles in the Lithuanian population for **A.** *HLA-A*, *HLA-B* and *HLA-C*. **B.** Nested distribution of guide RNAs on the gene body, exons, ectodomain and those with predicted high-activity for *HLA-A*, *HLA-B* and *HLA-C*. **C.** Protein structure models and gene structures for HLA-A, HLA-B and HLA-C. Protein structures are depicted as mature forms, excluding the signal peptide and without the highly flexible endodomain. Gene structure highlights the matching ectodomain and transmembrane (TM).

### Modeling the impact of HLA class I engineering on the immune compatibility of triple homozygous and double homozygous donor samples

Naturally occurring triple and double homozygous samples are particularly useful for gene engineering approaches as they allow bi-allelic targeting with a single programmable nuclease in a one-step intervention. Next, we modelled the impact of *HLA-A* and *HLA-B* knock-out on the immune compatibility of the double and triple homozygous samples when matching them to the Lithuanian population and pan-European datasets (**Figure 8**). We included all 51 triple homozygous individuals from our cohort (**Figure 8A**). From the 153 double homozygous individuals identified, we focused on those that are *HLA-A* and *HLA-B* double homozygous, comprising 7 individuals (**Figure 8B**). The 51 triple homozygous samples, when in an *HLA-C* retained (*HLA-A* and *HLA-B* double knock-out) configuration, match a maximum of 0.9799 of the Lithuanian population (**Figure 8A**). These 51 samples achieve a match of 0.9577 in the European-American population (**Figure 8C**) and 0.9556 on the British population (**Figure 8D**).

**Figure 8.**
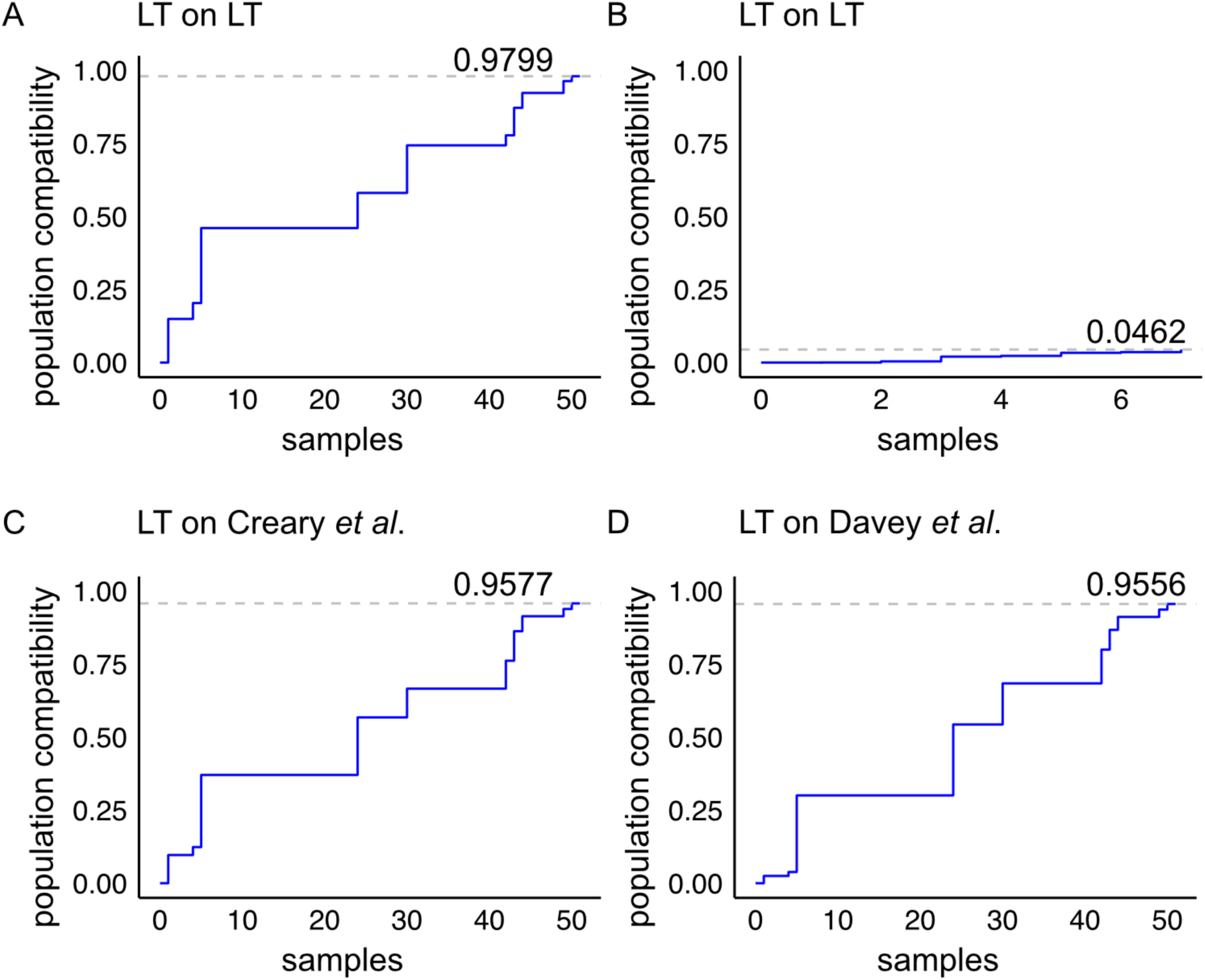
Population compatibility model of *HLA-A* and *HLA-B* double knock-out samples from our cohort with the Lithuanian and pan-European populations. **A.** Immune compatibility of the 51 triple homozygous individuals in an *HLA-A* and *HLA-B* double knock-out model, and **B.** the 7 double homozygous individuals in an *HLA-A* and *HLA-B* double knock-out model when matched to the Lithuanian population. Cohort from **A** when matched to the **C.** European-American dataset and **D.** the British dataset.

### Sampling of *HLA-A*, *HLA-B* and *HLA-C* triple homozygous individuals from the Lithuanian population

Since the triple and double homozygous individuals identified in this study are immune-compatible with a large fraction of the Lithuanian and pan-European population, we recruited these volunteer donors to collect dermal fibroblast primary samples for establishing biobank stocks and cultures. Primary fibroblast cultures were robustly established for 16 triple and double homozygous individuals (**Figure 9A**). Sanger sequencing of the PCR products of exon 2 and exon 3, which code for the ectodomain of *HLA-A*, *HLA-B*, and *HLA-C*, revealed characteristic residues for each allele. Characteristic amino acids p.F33 and p.R121 were confirmed for HLA-A*02:01:01:01 (**Figure 9C**), p.Y33 and p.W119 for HLA-B*13:02:01:01 (**Figure 9D**), and p.D33 and p.L119 for HLA-C*06:02:01:01 (**Figure 9E**). These findings were consistent for both the XY donor (donor SD9) and XX donor (donor SD6) individuals with homozygous haplotypes HLA-A*02:01:01:01-HLA-B*13:02:01:01-HLA-C*06:02:01:01 (**Figure 9F**).

**Figure 9:**
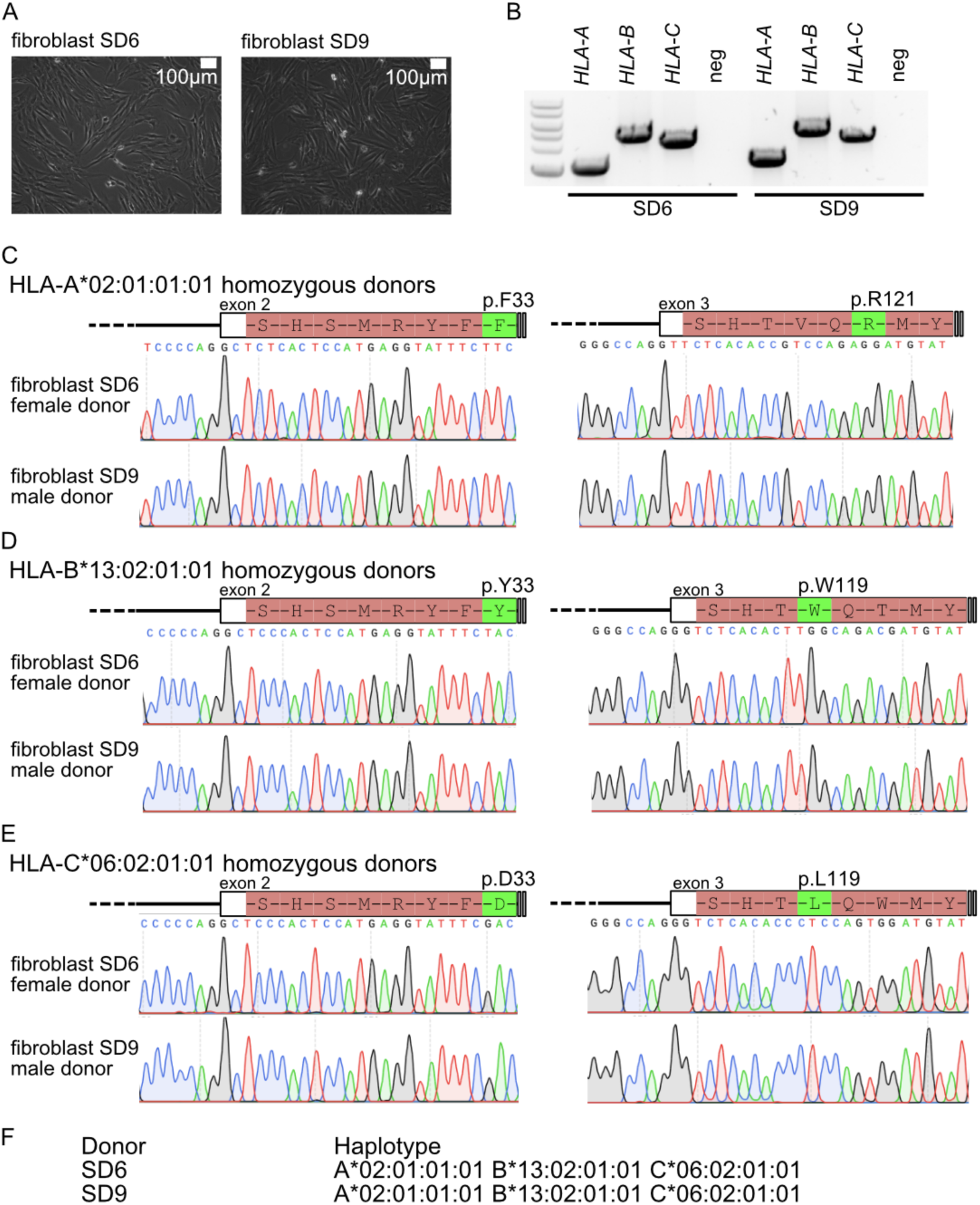
**A.** Fibroblasts cultures from *HLA-A*, *HLA-B* and *HLA-C* triple homozygous donors. **B.** Genotyping PCR for *HLA-A*, *HLA-B* and *HLA-C*. Sanger sequencing analysis for **C.** *HLA-A*, **D.** *HLA-B* and **E.** *HLA-C*. **F.** Next generation sequencing haplotype for donor patient and linked fibroblasts.

## 5. Discussion

Our study on allele and haplotype frequencies of the *HLA-A*, *HLA-B* and *HLA-C* genes in the Lithuanian population elucidates immune compatibility structure in relation to other European populations. Comparative analyses confirmed a high degree of similarity in HLA immune compatibility genes between the Lithuanian population and pan-European groups. The most frequent alleles described in the British ^20^ and European-American populations ^19^ are also the most frequent in the Lithuanian population, with frequencies of 31.7% (A*02:01:01), 5.3% (B*08:01:01), 15.0% (B*07:02:01), and 8.7% (C*07:01:01). Linear regression analysis using publicly available data corroborated these observations. Principal component analysis (PCA) and Euclidean distance calculations further confirmed the proximity in immune compatibility between Lithuanian, European-American, and British populations. Our Hardy-Weinberg equilibrium (HWE) analysis revealed deviations in a subset of alleles, suggesting partial genetic isolation or selective pressure. This finding aligns with previous studies indicating low levels of admixture and a significant component of pre-Neolithic hunter-gatherer ancestry in the Lithuanian group ^18^.

The majority of individuals in our registry (n = 11,153) were characterized at second-field resolution for *HLA-A*, *HLA-B*, and *HLA-C*, while a subset (n = 3,496) underwent third-field resolution analysis. This divergence reflects technological advancements in clinical registries, with long-read sequencing platforms now enabling fourth-field resolution ^27,28^. Although our analyses do not encompass HLA class II, it is well established that its expression occurs in specialized immune cell lineages, while HLA class I primarily regulates non-immune cell compatibility. Remarkably we found a subset of 51 triple homozygous for *HLA-A*, *HLA-B* and *HLA-C*, and a subset of 153 double homozygous individuals. The proportion of triple-homozygous individuals exceeded stochastic expectations based on measured allele frequencies (2.99 ± 1.76), suggesting underlying population structures, as indicated by HWE analysis.

Due to the significant immune compatibility provided by *HLA-A*, *HLA-B* and *HLA-C* triple homozygous individuals ^29,30^ the term naturally-occurring “*super donors*” has been proposed ^31^. Our study identified 51 naturally-occurring super donors, who exhibit class-I immune matching with 60.54% of the Lithuanian population, 13.4% of the British population, and 7.4% of the European-American population. It is important to highlight that using triple homozygous samples for cell line development, particularly human induced pluripotent stem (iPS) cells, results in derivatives with wider immune compatibility than heterozygous counterparts. Genetic engineering with programmable nucleases in such samples benefits from simpler strategies, because of the homozygosity status of the starting material. In turn, engineered products are expected to attain broader immune compatibility than natural counterparts.

Several international initiatives focus on iPSC development from haplo-selected individuals, including programs in Japan ^32^, Australia ^31^, South Korea ^33,34^, Spain ^35^, Germany ^36^, Lithuania, Saudi Arabia ^37^. We modeled the impact of “*HLA-C retained*” gene-editing intervention on the 51 naturally occurring *super donors* and found that their immune compatibility could be enhanced to match 97.9% of the Lithuanian population, 95.7% of the European-American population and 95.5% of the British population. Conversely, the immune compatibility provided by the *HLA-A* and *HLA-B* double-homozygous was limited due to the retained diversity within the heterozygous HLA-C allele.

Here, we propose the term “*synthetic superdonor”* for those cell lines derived from naturally occurring *superdonors* that, through means of gene editing, acquire broader immune compatibility. Analysis on the gene editing availability for *HLA-A*, *HLA-B* and *HLA-C* highlights the importance of protein topology, knock-out strategy design and nuclease target site activity to achieve *synthetic superdonor* stocks. The HLA-A, HLA-B and HLA-C proteins are of the type-I transmembrane class; hence, targeting the N-terminus ectodomain slightly constrains the number of available Cas9 binding sites. Our analyses demonstrate that the largest impact to knock-out availability is the nuclease activity score; therefore, gene editing tools that enhance nuclease activity are likely to have a positive impact on *synthetic superdonor* creation in the future. Likewise, our analyses indicate that naturally occurring *superdonor* and *synthetic superdonor* cell sources would positively impact the immune matching for rare haplotypes. Both, naturally occurring and *synthetic superdonors* offer are a remarkable source for regenerative medicine applications and for the creation of derivative advanced therapeutic medicinal products (ATMPs).

## Acknowledgements

This project was supported by Mission-driven Implementation of Science and Innovation Program 02-002-P-0001 to D.B. and J.A., funded by the Economic Revitalization and Resilience Enhancement Plan “New Generation Lithuania”. All authors read and agreed to the last version of the manuscript.

## Disclosures

The authors of this manuscript have no conflicts of interest to disclose.

## Data availability statement

All data is made available through public repositories.

## Notes

### Competing Interest Statement

The authors have declared no competing interest.

### Author Declarations

Ethical approval. This study is part of the ethical approval 2023/6-1524-984 - Highly-immune compatible iPS cells as source for Regenerative Medicine and Cell Therapy-oriented applications - from the Vilnius Regional Biomedical Research Ethics Committee to Vilnius University and 2023/4-1507-968 - Analysis of the Distribution of Human Leukocyte Antigen (HLA Encoding Genes - HLA) Alleles and Haplotypes in the Group of the Lithuanian Unrelated Bone Marrow Donor Registry - to Vilnius University Hospital Santaros Klinikos. Written consents were obtained from the participants of the study.

